# How does the risk of bias influence the effect sizes of exercise therapy in chronic low back pain randomised controlled trials? A protocol for a meta-epidemiological study

**DOI:** 10.1101/2022.07.20.22277703

**Authors:** Tiziano Innocenti, Jill A Hayden, Stefano Salvioli, Silvia Giagio, Leonardo Piano, Carola Cosentino, Fabrizio Brindisino, Daniel Feller, Rachel Ogilvie, Silvia Gianola, Greta Castellini, Silvia Bargeri, Jos WR Twisk, Raymond Ostelo, Alessandro Chiarotto

## Abstract

**Background and Objectives:** Risk of bias is a critical issue to consider when appraising studies. Generally, the higher the risk of bias of a study, the less confidence there will be that the results are valid. Considering that low back pain is recognized to have an extremely high disease burden; exercise therapy is one of the most frequently prescribed interventions for chronic low back pain (CLBP) and that most low back pain trials have methodological limitations that could bias treatment effect estimates; the objective of this study is to explore causal pathways between the sources of risk of bias and estimates of the treatment effect of exercise therapy interventions in CLBP trials.

**Methods:** The 249 RCTs included in the 2021 Cochrane review publication “Exercise therapy for chronic low back pain” will be included. The risk of bias will be evaluated with the Cochrane Risk of Bias 2 tool (ROB 2). Causal pathways between the exposure (risk of bias domains) and our outcomes of interest (effect sizes for pain and functional limitations) will be explored through univariable and multivariable meta-regression models. These models will be adjusted for potential confounders (sample size, trial registration, incomplete flow chart information and treatment comparisons), exploring relevant interactions within each model. Additional and sensitivity analyses will be performed to explore and test the robustness of the primary analyses.

**Ethics and dissemination:** A manuscript will be prepared and submitted for publication in an appropriate peer-reviewed journal upon study completion. We believe that the results of this investigation will be relevant to researchers paying more attention to the synthesis of the evidence to translate clinical implications to key stakeholders (healthcare providers and patients).

## INTRODUCTION

Risk of bias is a critical issue to consider when appraising studies. Generally, the higher the risk of bias of a study, the less confidence there will be that the results are valid. Clinicians, who are the primary consumers of the evidence generated by randomized controlled trials (RCTs), would benefit from a clear understanding of the potential for biased results before incorporating findings from the trials into clinical practice. Previous meta-research studies^1,2^ demonstrated that treatment effect estimates were overestimated in trials with specific methodological limitations (e.g. inadequate allocation concealment and lack of double-blinding). Recently, other studies have investigated the potential influence of specific factors on effect sizes at the study level^3-5^. For example, Faggion et al.^5^ evaluated the influence of three characteristics (i.e. sequence generation, allocation concealment, and blinding of outcome assessment) on the size of meta-analytic estimates in systematic reviews of RCTs published in periodontology and implant dentistry, finding that high and unclear risks of selection and detection biases did not appear to influence the size of treatment effect estimates; however, several confounders may have influenced the strength of the association. Most meta-research studies derived from reports of RCTs are from the broad field of medicine.

In the physical therapy field, few meta-epidemiological studies have been published, most of them focused on chronic non-specific low back pain (CLBP). This is because CLBP is the most prevalent, costly, and disabling musculoskeletal condition with the largest number of clinical trials in the physical therapy literature^6^. However, most of these studies investigated only specific characteristics of the risk of bias domains. For example, Armijo-Olivo et al.^7^ investigated the influence of sequence generation and allocation concealment on treatment effects. They did not find an association between these variables and treatment effects, but they included studies from different areas of physical therapy, selecting a wide range of different clinical conditions with significant heterogeneity that may have influenced this possible association. Oliveira de Almeida et al.^8^ tried to overcome this issue by focusing on physical therapy interventions in low back pain, investigating only the influence of allocation concealment and intention-to-treat analysis on the effect estimates by using the PEDro scale. They found no influence of allocation concealment or intention-to-treat analysis on treatment effects of physical therapy interventions for pain and disability in low back pain trials.

Recently, a meta-epidemiological study led by Hayden et al.^9^ investigated the association of research integrity characteristics (publication integrity, trial registration, high ROB, sample size and other features at the study level) and the effect size in a subgroup of RCTs included in a Cochrane review^10^ on exercise therapy for CLBP published in 2021. The authors found an association between some study characteristics (such as sample size and the use of core outcome set) and the effect sizes.

Considering that CLBP is recognized to have an extremely high disease burden;^11^ exercise therapy is one of the most frequently prescribed interventions for this condition^12^ and that most low back pain trials have methodological limitations that could bias treatment effect estimates;^13,14^ the objective of this study is to explore causal pathways between the sources of risk of bias and estimates of the treatment effect of exercise therapy interventions in CLBP trials.

## MATERIAL AND METHODS

We will follow the adaptation of the PRISMA 2009 for meta-epidemiological studies proposed by Murad et al.^15^ for the reporting of this manuscript (or a specific reporting checklist for meta-epidemiological studies, if available at the time of reporting^16^).

### CAUSAL PATHWAY

Despite the growing number of studies published every year, one of the most common recognized issues that affect meta-epidemiological research is the failure to explicitly acknowledge a causal objective and the lack of a gold-standard approach to analysis and control for potential sources of confounding^17,18^.

Considering our objective, we need to control for a selection of potential confounders that might influence the relationship between the sources of bias (exposures) and the observed effect sizes (outcomes of interest). To do that, we have drawn a directed acyclic graph (DAG) to make explicit our hypotheses about the network of important causes of the observed effect of the intervention (see Figure 1 in Appendix 1). The DAG was drawn and will be analyzed using DAGitty software^19^.

Sample size, incomplete information about the flow of the participants through the trial (i.e., flow chart), and the study’s registration status (prospective, retrospective, unregistered) are variables that may influence both the risk of bias assessment and the effect size. In the recent study by Hayden et al.^9^ these variables showed a trend of association with the effect size in a similar subset of low back pain trials. Another source of potential confounding is the comparison intervention: it is well established that the type of comparison influences both the effect sizes^4,17^ and the risk of bias.

Regarding the biases in the measurement of outcome, for instance, when the outcome assessor is not blinded, and the outcome may be influenced by knowledge of the intervention received, there could be a higher risk of bias; this is especially true if the comparator is no treatment or usual care. When the comparator is another active intervention, outcome assessors (and participants) may not have a prior belief that one of the active interventions is more beneficial than the other. Other potential confounders, such as the disease and the type/construct of outcomes, are controlled by our selection criteria.

### DATA SOURCE

The 249 RCTs included in the 2021 Cochrane review publication “Exercise therapy for chronic low back pain”^10^ will be included in this study. The review evaluated the efficacy of exercise therapy interventions in adults with CLBP; they assessed pain intensity and functional limitations (as continuous variables) as their primary outcomes. The dataset from the study conducted by Hayden et al.^9^ will also be used to extract relevant data.

### DATA EXTRACTION

#### Descriptive data

We will access descriptive study data previously extracted for the original Cochrane review^10^, including study design characteristics, setting, year, population, interventions, comparisons, and outcome information.

These data were recorded on pre-developed and tested forms using web-based electronic systematic review software (Distiller SR. Ottawa, ON, Canada: Evidence Partners, 2020). A single reviewer extracted study information, and at least one other author checked all extracted points against the original study publications. For further details about the data extraction, a detailed explanation is provided in the original publication^10^.

#### Covariates

The following variables will be used as potential confounders and/or effect modifiers:

⍰ Sample size (continuous)
⍰ Trial registration (if the included RCTs were prospectively registered and/or a complete study protocol is available) (dichotomous – yes; if the study has been prospectively registered/no; if the study has been retrospectively registered or there is no registration or published protocol)
⍰ Incomplete flow chart information (dichotomous – yes/no)
⍰ Comparisons as follows (categorical):
  - Exercise vs Placebo
  - Exercise vs No intervention (including usual/normal care, waitlist or same co-interventions)
  - Exercise vs Another intervention
  - Exercise as an add-on intervention vs another intervention alone

#### Risk of Bias

As recommended by the Cochrane Handbook, we will use the most recent Cochrane risk of bias tool, the Cochrane Risk of Bias 2 Tool^20^ (RoB 2 – 22 August 2019 version) to assess the risk of bias in included trials. The evaluation will be done using pre-tested, web-based forms using DistillerSR software^21^. The risk of bias will be assessed for each outcome separately – pain intensity and functional limitations (our outcomes of interest and the most relevant core domains in low back pain^22^). For each outcome, a risk of bias rating will be calculated for each of the five domains using the algorithm set out by the RoB 2 developers (see the full RoB 2 guidance document for a detailed description of each domain and assessor guidance materials^23^):

⍰ Domain 1: bias arising from the randomisation process
⍰ Domain 2: bias due to deviations from intended interventions
⍰ Domain 3: bias due to missing outcome data
⍰ Domain 4: bias in the measurement of the outcome
⍰ Domain 5: bias in the selection of the reported result

The risk of bias ratings for domains within the RoB 2 tool are ‘‘high,’’ ‘‘low,’’ or ‘‘some concerns’’. The overall risk of bias score will not be extracted.

The risk of bias evaluation will be performed considering the following criteria (to be aligned with the guidance of the RoB 2 tool):

⍰ We will only extract outcome results for the short-term follow-up (closest to 12 weeks).
⍰ When an outcome is evaluated in a trial using more than one scale, we will prioritize according to the following order^24,25^:
  ⍰ **Pain:**
    1. numerical rating scale (NRS)
    2. visual analogue scale (VAS)
    3. Pain severity subscale of the Brief Pain Inventory (BPI)
    4. Any other scale
  ⍰ **Functional Limitations:**
    1. Roland-Morris Disability Questionnaire (RMDQ)
    2. Oswestry Disability Index (ODI)
    3. Quebec Back Pain Disability Scale (QBPDS)
    4. Any further back pain-specific scale
⍰ When an outcome measure is recorded more than once during a relevant follow-up period (e.g., NRS during last week, NRS last 24 hours, NRS at rest, etc.), we will choose one according to the following preference order:
  - During last week
  - During the last 24 hours
  - At the moment (1. at rest, 2. At night, 3. Anything else)
  - Any other timeframe
⍰ We will only extract outcome results for the short-term follow-up (closest to 12 weeks). When the trial has more than one parallel arm, we will choose the comparison between exercise therapy and one of the following according to this preference order:
  1. No treatment, usual care or placebo
  2. Other conservative therapy
  3. Another exercise group
⍰ The effect of interest will be the effect of assignment to intervention (“intention to treat” effect as indicated in the RoB 2).

Five pairs of reviewers with prior experience using the tool will conduct the risk of bias assessments independently; disagreements will be resolved through consensus within the pairs, with a third reviewer consulting when required to reach an agreement.

##### Calibration Phase

To improve inter-rater reliability, all team members participated in a two-stage calibration exercise facilitated by the Exercise for chronic low back pain Network Review (forthcoming, Hayden et al., 2022) central team in DistillerSR^21^. All team members were required to complete this calibration exercise before the assessment of RoB 2 for included trials. In the first stage of calibration, team members were divided into two-person teams. Each team was required to conduct a risk of bias assessment and reach a consensus for four purposively selected trials. These four trials were chosen to include common and less common but challenging bias-related issues. In the second calibration stage, all teams were provided with a document comparing their individual and consensus responses to those of two Network Review central team members for review and discussion. In addition, the project leader (TI) was provided with a set of common issues and areas of divergence identified across the whole team. As the final component of calibration, TI held a debrief meeting with all team members to reach a consensus on consistent approaches to address areas of divergence.

## DATA ANALYSIS

### Descriptive Analysis

We will report study design characteristics, setting, year, population, interventions, comparisons, and outcome information. We will descriptively report the results, displaying the risk of bias rating for each RoB 2 domain using traffic light plots.

### Statistical Analysis

Considering that all studies assess the same outcomes (pain and/or functional limitations) but measure them in various ways, we will use the standardised mean difference (SMD).

We will construct both univariable and multivariable meta-regression models considering our study hypothesis. The dependent variables will be the treatment effect estimates (SMD between groups) for the outcomes of interest (pain intensity and functional limitations separately). In the primary analyses, the independent variables will be the risk of bias (categorised as a dichotomous variable “Low vs High/Some concerns”) in each RoB 2 domain.

#### Univariable models

For each univariable model, we will perform two different analyses:

1. Unadjusted
2. Adjusted for potential confounders

##### 1) Unadjusted analysis

We will perform five meta-regression analyses separately for each outcome (pain intensity and functional limitations), in which the dependent variable will be the treatment effect (SMD between groups). The independent variables will be the risk of bias (categorised as a dichotomous variable “Low vs High/Some concerns”) in each RoB 2 domain.

These models are summarized in Table 1.

**Table 1:** Unadjusted univariable models.

| Meta-regression analysis | Dependent variable | Independent variables |
| --- | --- | --- |
| Univariable Model 1 | Effect estimates<br>☑ Pain intensity<br>☑ Functional limitations | ☑ RoB domain 1: Low vs. High/Some concerns |
| Univariable Model 2 |  | ☑ RoB domain 2: Low vs. High/Some concerns |
| Univariable Model 3 |  | ☑ RoB domain 3: Low vs. High/Some concerns |
| Univariable Model 4 |  | ☑ RoB domain 4: Low vs. High/Some concerns |
| Univariable Model 5 |  | ☑ RoB domain 5: Low vs. High/Some concerns |

##### 2) Adjusted analysis

The following covariates will be added in the adjusted analyses to the models mentioned above depending on their role as potential confounders (i.e., we will use in each model the variable that we have identified in the DAG as potential confounders):

⍰ Sample size (continuous data)
⍰ Trial registration (if the included RCTs have been registered and a complete study protocol is available) (dichotomous – yes/no)
⍰ Incomplete flow chart information (dichotomous – yes/no)
⍰ Treatment comparisons (dummy variables with exercise as the reference)
  - Exercise vs Placebo
  - Exercise vs No intervention (including usual/normal care, waitlist or same co-interventions)
  - Exercise vs Another intervention
  - Exercise as an add-on intervention vs another intervention alone

These models are summarized in Table 2

**Table 2:** Adjusted univariable models.

| Meta-regression analysis | Dependent variable | Independent variables | Adjustment variables |
| --- | --- | --- | --- |
| Univariable Model 1 | Effect estimates<br>☐ Pain intensity<br>☐ Functional limitations | ☐ RoB domain 1: Low vs. High/Some concerns | No confounders were identified |
| Univariable Model 2 | Effect estimates<br>☐ Pain intensity<br>☐ Functional limitations | ☐ RoB domain 2: Low vs. High/Some concerns | ☐ Sample size<br>☐ Incomplete flow chart information<br>☐ Treatment comparisons:<br>○ Exercise vs Placebo<br>○ Exercise vs No intervention<br>○ Exercise vs Another intervention<br>○ Exercise as an add-on intervention vs another intervention alone |
| Univariable Model 3 | Effect estimates<br>☐ Pain intensity<br>☐ Functional limitations | ☐ RoB domain 3: Low vs. High/Some concerns | ☐ Sample size<br>☐ Incomplete flow chart information<br>☐ Treatment comparisons:<br>○ Exercise vs Placebo<br>○ Exercise vs No intervention<br>○ Exercise vs Another intervention<br>○ Exercise as an add-on intervention vs another intervention alone |
| Univariable Model 4 | Effect estimates<br>☐ Pain intensity<br>☐ Functional limitations | ☐ RoB domain 4: Low vs. High/Some concerns | ☐ Trial registration<br>☐ Treatment comparisons:<br>○ Exercise vs Placebo<br>○ Exercise vs No intervention<br>○ Exercise vs Another intervention<br>○ Exercise as an add-on intervention vs another intervention alone |
| Univariable Model 5 | Effect estimates<br>☒ Pain intensity<br>☒ Functional limitations | ☒ RoB domain 5: Low vs. High/Some concerns | ☒ Trial registration |

#### Multivariable model

We will perform a multivariable model for each outcome (pain intensity and functional limitations), in which the dependent variable will be the treatment effect (SMD between-group). The independent variables will be the risk of bias (categorised as dummy variables “high vs some concerns” and “high vs low”) in each RoB 2 domain.

This model is summarized in Table 3.

**Table 3:** Unadjusted multivariable model(s)

| Meta-regression analysis | Dependent variable | Independent variables |
| --- | --- | --- |
| Multivariable Model(s) | Effect estimates:<br>☐ Pain intensity<br>☐ Functional limitations | ☐ RoB domain 1: Low vs. High/Some concerns<br>☐ RoB domain 2: Low vs. High/Some concerns<br>☐ RoB domain 3: Low vs. High/Some concerns<br>☐ RoB domain 4: Low vs. High/Some concerns<br>☐ RoB domain 5: Low vs. High/Some concerns |

As we have described above, also for the multivariable model, we will adjust for the covariates (Table 4).

**Table 4:** Adjusted multivariable model(s)

| Meta-regression analysis | Dependent variable | Independent variables | Adjustment variables |
| --- | --- | --- | --- |
| Multivariable Model(s) | Effect estimates<br>☐ Pain intensity<br>☐ Functional limitation | ☐ RoB domain 1: Low vs. High/Some concerns<br>☐ RoB domain 2: Low vs. High/Some concerns<br>☐ RoB domain 3: Low vs. High/Some concerns<br>☐ RoB domain 4: Low vs. High/Some concerns<br>☐ RoB domain 5: Low vs. High/Some concerns | ☐ Sample size<br>☐ Incomplete flow chart information<br>☐ Trial registration<br>☐ Treatment comparisons:<br>○ Exercise vs Placebo<br>○ Exercise vs No intervention<br>○ Exercise vs Another intervention<br>○ Exercise as an add-on intervention vs another intervention alone |

For each linear regression model, the assumptions of linearity, homoscedasticity, independence and normality will be checked. All analyses will be performed using SPSS software (IBM SPSS Statistics for Macintosh, Version 28.0. Armonk, NY: IBM Corp).

## ADDITIONAL ANALYSES

We will perform additional analyses to explore and test the robustness of the primary analyses. We will replicate the analyses using the risk of bias categorised as dummy variables with low risk as reference (“Low vs High” and “Low vs Some concerns”).

### Sensitivity Analyses

We will conduct the following sensitivity analyses:

1. limiting to studies published in 2010 or later to account for good dissemination of reporting guidelines and standards (i.e. publication data of the CONSORT checklist^26^).
2. limiting to studies published in 2007 or later, that is two years after the International Committee of Medical Journal Editors (ICMJE) ‘obligation’ to report^27^.

## Data Availability

All data produced in the present study are available upon reasonable request to the authors

## ETHICS AND DISSEMINATION

This study does not require ethics review as we will not be collecting personal data; it will summarise information from publicly available studies.

A manuscript will be prepared and submitted for publication in an appropriate peer-reviewed journal upon study completion. The study findings will be disseminated at a relevant (inter)national conference. All results of this meta-epidemiological study will also be announced at (inter)national scientific events in musculoskeletal rehabilitation and research methods. We believe that the results of this investigation will be relevant to researchers paying more attention to the synthesis of the evidence to translate clinical implications to key stakeholders (healthcare providers and patients).

## AUTHOR CONTRIBUTIONS

TI, AC, JH, and RO conceived and designed the study protocol. TI, JA, SS, SG, LP, CC, FB, DF, RO, SG, GC, SB, JWRT, RO and AC were involved in conceptualising the study objectives, providing input into study selection criteria and plans for data extraction. All the authors, including TI, JA, SS, SG, LP, CC, FB, DF, RO, SG, GC, SB, JWRT, RO and AC, approved the final version of the protocol.

## FUNDING STATEMENT

This research received no specific grant from any funding agency in the public, commercial or not-for-profit sectors. SG, GC and SB were supported by the Italian Ministry of Health “Linea 2 – Studi metodologici in ortopedia e riabilitazione”-L2085.

---

**Appendix 1 – Figure 1:**
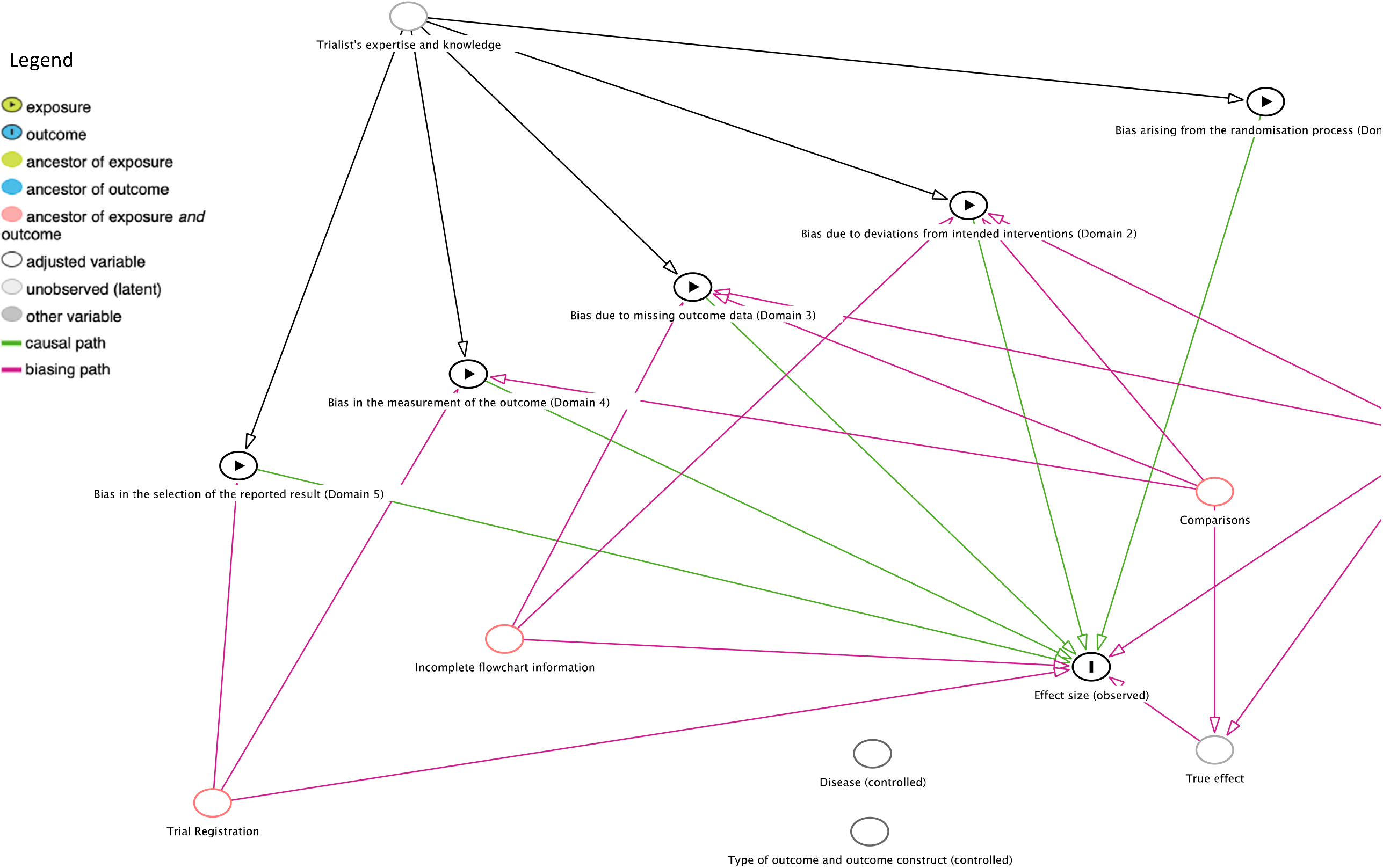
A directed acyclic graph (DAG) formalizing the author’s beliefs about the network of causes surrounding estimates of the effects of intervention in randomized controlled trials.

## Notes

### Competing Interest Statement

The authors have declared no competing interest.

### Funding Statement

This research received no specific grant from any funding agency in the public, commercial or not-for-profit sectors. Silvia Gianola, Greta Castellini and Silvia Bargeri were supported by the Italian Ministry of Health Linea 2 Studi metodologici in ortopedia e riabilitazione; L2085.

### Summary of Updates

We removed the following part from the analyses paragraph: 3) Exploring relevant interactions Lastly, we will investigate whether relevant interactions could exist between the covariates and the risk of bias domains. When significant interactions were found, separate results will be reported for different values of the effect modifier. Rationale: We decided to leave the interactions out of the multivariable model and only perform a crude and adjusted analysis to be more aligned with our hypothesis. That change was done before the start of the data analysis. If a specific reporting checklist for meta-epidemiological studies will be not available at the time of reporting, we will follow the adaptation of the PRISMA 2009 for meta-epidemiological studies proposed by Murad et al. (doi: 10.1136/ebmed-2017-110713)

